# Inflammation Proteomic Profiling of Psychosis in Young Adults: findings from the ALSPAC birth cohort

**DOI:** 10.1101/2024.04.29.24306568

**Authors:** Ruby S. M. Tsang, Nicholas J. Timpson, Golam M. Khandaker

**Author notes:** Corresponding author: Ruby Tsang, Population Health Sciences, Bristol Medical School Oakfield House, Oakfield Grove Bristol BS8 2BN United Kingdom. Declaration of Interest: None.

## Abstract

Psychotic disorder is associated with altered levels of various inflammatory markers in blood, but existing studies have typically focused on a few selected biomarkers, have not examined specific symptom domains notably negative symptoms, and are based on individuals with established/chronic illness. Based on data from young people aged 24 years from the Avon Longitudinal Study of Parents and Children (ALSPAC), a UK birth cohort, we have examined the associations of 67 plasma immune/inflammatory proteins assayed using the Olink Target 96 Inflammation panel with psychotic disorder, positive (any psychotic experiences and definite psychotic experiences) and negative symptoms, using linear models with empirical Bayes estimation. The analyses included between 2641 and 2854 individuals. After adjustment for age, sex, body mass index and smoking and correction for multiple testing, upregulation of CDCP1 and IL-6 were consistently associated with positive symptoms and psychotic disorder, while psychotic disorder was additionally associated with upregulation of MMP-10. Negative symptoms were associated with upregulation of the highest number of proteins (n=11), including cytokines, chemokines and growth factors which partly overlap with proteins associated with positive symptoms or psychotic disorder (CDCP1, IL-6 and MMP-10). Our findings highlight associations of inflammatory proteins involved in immune regulation, immune cell activation/migration, blood-brain barrier disruption, and extracellular matrix abnormalities with psychosis or psychotic symptoms in young people, consistent with a role of inflammation and immune dysfunction in the pathogenesis of psychotic disorders.

## 1. Introduction

Psychosis is an umbrella term that refers to a group of complex neuropsychiatric disorders primarily characterized by positive (e.g., hallucinations, delusions, or confusion) and negative symptoms (e.g., apathy and diminished expression). The median lifetime prevalence for schizophrenia, the archetypal psychotic disorder, is 4.0 per 1000 persons (McGrath et al., 2008), whereas it is 7.5 per 1000 persons for all psychoses (Moreno-Küstner et al., 2018). In contrast, subthreshold psychosis-like experiences are relatively common and are experienced by approximately a quarter of the general population during their lifetime (Bourgin et al., 2020). It has been suggested that the psychosis phenotype is expressed on a continuum, where symptoms may present in non-clinical populations (van Os et al., 2009).

Schizophrenia is a multifactorial condition with both genetic and environmental contributions. Infection, inflammation and immune dysfunction have been implicated in the pathogenesis of schizophrenia and related psychoses (Benros et al., 2011; Brown and Derkits, 2010; Goldsmith et al., 2016; Khandaker et al., 2015). There is substantial evidence for cross-sectional associations between central and peripheral inflammation and schizophrenia spectrum disorders. Abnormal cerebrospinal fluid (CSF) markers were observed in individuals with schizophrenia spectrum disorders, including elevated total protein, albumin ratio, white blood cell count, glucose, IL-6 and IL-8 levels (Warren et al., 2024). Blood concentrations of a number of inflammatory markers, namely interleukin (IL)-1β, IL-1 receptor antagonist (IL-1RA), soluble IL-2 receptor (sIL-2R), IL-6, IL-8, IL-10, tumor necrosis factor (TNF)-α, and C-reactive protein (CRP) were consistently elevated in individuals with schizophrenia-spectrum disorder compared to healthy controls (Halstead et al., 2023).

Subsets of cytokines may be associated with specific features of the disorder, for instance IL-2 and interferon (IFN)-γ were elevated in acute schizophrenia-spectrum disorder whereas IL-4, IL-12 and IFN-γ were reduced in chronic schizophrenia-spectrum disorder (Halstead et al., 2023); another study found elevated IL-6, IL-12, IL-17 and IFN-γ levels were observed in antipsychotic-naïve first-episode psychosis (FEP) while elevated IL-1β, IL-2, IL-4, IL-6 and TNF-α as well as reduced IL-10, were associated with negative symptoms (Dunleavy et al., 2022). Increased total white blood cell, monocyte and neutrophil counts were also observed in schizophrenia and FEP (Jackson and Miller, 2020). Similar dysregulations of inflammatory markers are also seen in those with subthreshold or transitory symptoms; when comparing subgroups with familial risk and clinical risk of psychosis with controls, the clinical risk subgroup showed higher IL-6 levels than controls, but the study did not find any evidence for markers that predicted transition to psychosis (Misiak et al., 2021).

Longitudinally, exposures to inflammation during gestation and in childhood have been shown to be associated with psychosis risk. Prenatal exposure to infections and elevated proinflammatory cytokines may be associated with schizophrenia risk and structural and functional brain abnormalities relevant to schizophrenia, though effects may differ by infectious agent and timing of exposure (Khandaker et al., 2013). Furthermore, childhood viral infections involving the central nervous system may be associated with an increased risk of adult schizophrenia (Khandaker et al., 2012). Studies using data from the Avon Longitudinal Study of Parents and Children (ALSPAC) also showed that circulating IL-6 and CRP levels assessed in childhood and adolescence predicted psychosis or schizophrenia risk in adulthood (Khandaker et al., 2014; Metcalf et al., 2017), suggesting that immune-inflammatory changes predate the onset of psychosis or schizophrenia. Two other studies using ALSPAC data compared protein expression levels in childhood between those who did and did not later develop psychotic experiences or disorder in early adulthood, and found dysregulation of the complement and coagulation system was associated with psychosis risk (English et al., 2017; Föcking et al., 2021).

Genome-wide association studies have identified multiple loci associated with schizophrenia risk, with strongest effects seen from variations in the major histocompatibility complex locus and particularly in complement component 4 genes, implicating immune function and complement activity in the pathogenesis of schizophrenia (Sekar et al., 2016). Furthermore, Mendelian randomization studies support a potentially causal role of IL-6 signaling in schizophrenia, with increased levels of IL-6 and soluble IL-6 receptor (sIL-6R) associated with a higher risk of schizophrenia; additionally, IL-6 appeared to explain the previously reported protective effect of CRP (Hartwig et al., 2017; Perry et al., 2021).

While existing studies support a role of inflammation in psychosis, there are key gaps in knowledge. For instance, most existing studies have typically focused on a limited number of inflammatory markers, but examining a larger immune proteomic panel can yield greater system-level insights into the pathophysiology of psychosis. Negative symptoms are core to schizophrenia, but there are relatively fewer studies of inflammation and negative symptoms compared to that of positive symptoms (Dunleavy et al., 2022). Negative symptoms represent an unmet clinical need as existing antipsychotic drugs do not adequately improve these symptoms. There is some evidence that inflammatory markers are associated with negative symptoms (Dunleavy et al., 2022; Khandaker et al., 2021); therefore, further studies are needed to examine these associations to understand whether inflammation could be a treatment target for negative symptoms. Finally, most existing studies are based on adults with established psychotic disorder. Studies of people who are relatively early on in the course of illness are required to identify immunological factors that are potentially relevant for incidence, rather than chronicity/prognosis. Studies of young people are also advantageous as this group is relatively unaffected by confounding from chronic inflammation-related diseases of adulthood.

We present a study examining the associations of a range of inflammatory proteins, assessed using a proteomic panel, with psychotic disorder and positive and negative symptoms of psychosis at age 24 years using data from the Avon Longitudinal Study of Parents and Children (ALSPAC), a UK birth cohort. The aim of this study was to examine the plasma inflammatory proteomic profile of psychotic disorder, and to compare patterns of associations for positive and negative symptoms in young adults.

## 2. Materials and Methods

### 2.1. Cohort and sample selection

Pregnant women resident in Avon, United Kingdom with expected dates of delivery between 1^st^ April 1991 and 31^st^ December 1992 were invited to take part in the Avon Longitudinal Study of Parents and Children (ALSPAC) (Boyd et al., 2012; Fraser et al., 2012; Northstone et al., 2019). The initial number of pregnancies enrolled was 14,541 and 13,988 children were alive at 1 year of age. Additional recruitment took place when the oldest children were approximately 7 years of age. The total sample size for analyses using any data collected after the age of seven is therefore 15,447 pregnancies, with 14,901 children who were alive at 1 year of age (Generation 1).

The present study focuses on the subset of Generation 1 participants who had data available for the relevant exposures (psychotic experiences, psychotic disorder, or negative symptoms), outcomes (blood immunological protein levels), and covariables (age, sex at birth, body mass index [BMI], and smoking). The sample selection flow diagram is presented in **Figure 1**.

**Figure 1.**
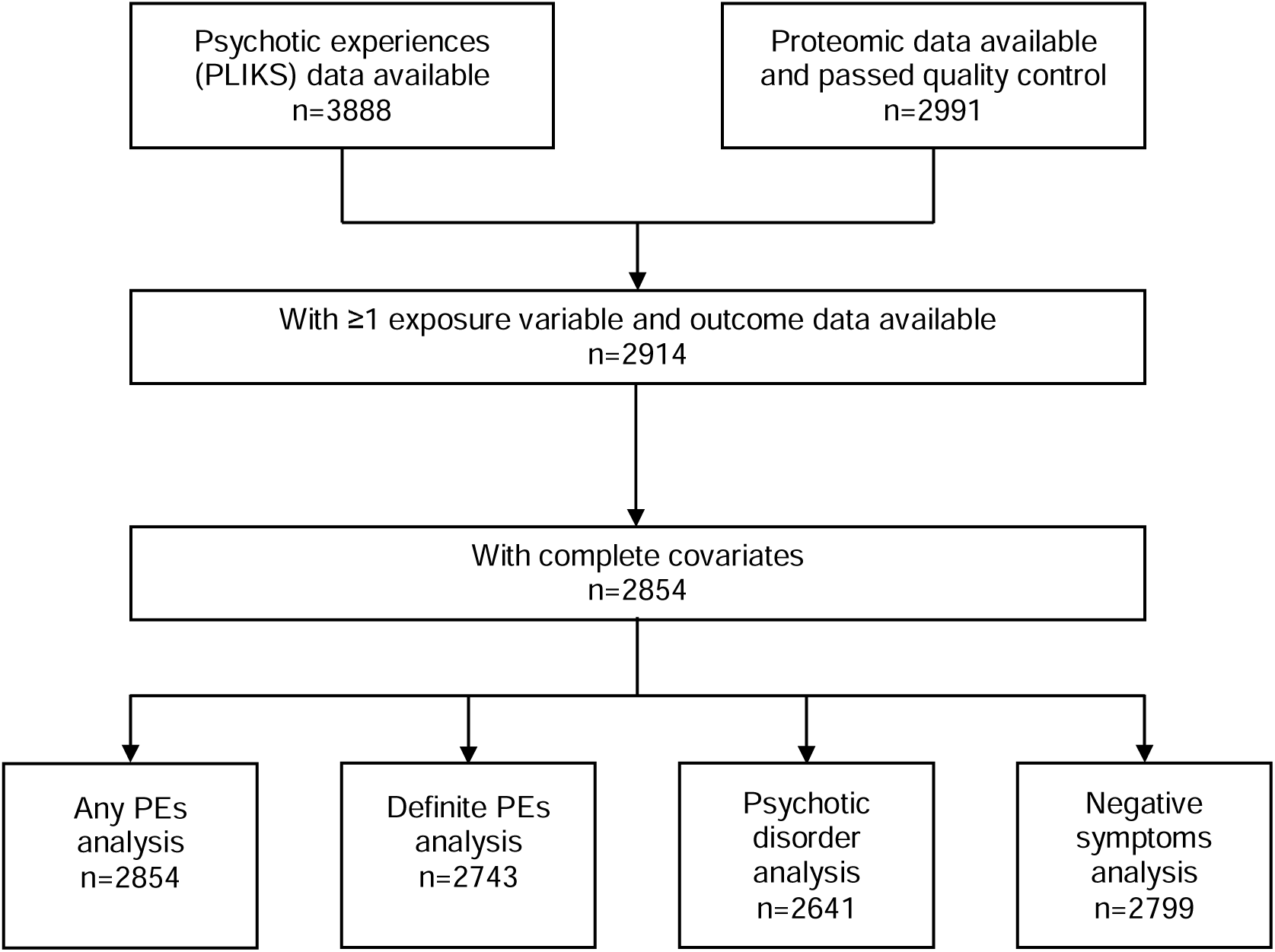
Flow diagram showing data availability in Generation 1 of ALSPAC for the analyses conducted.

Study data were collected and managed using Research Electronic Data Capture (REDCap) electronic data capture tools hosted at the University of Bristol (Harris et al., 2009). REDCap is a secure, web-based software platform designed to support data capture for research studies. Please note that the study website contains details of all the data that is available through a fully searchable data dictionary and variable search tool (http://www.bristol.ac.uk/alspac/researchers/our-data/).

### 2.2. Ethics

Ethical approval for the study was obtained from the ALSPAC Ethics and Law Committee and the Local Research Ethics Committees. Consent for biological samples has been collected in accordance with the Human Tissue Act (2004). Informed consent for the use of data collected via questionnaires and clinics was obtained from participants following the recommendations of the ALSPAC Ethics and Law Committee at the time.

### 2.3. Psychosis-related measures at age 24

Psychotic experiences (PEs) and disorder were identified through the face-to-face, semi-structured Psychosis-Like Symptoms Interview (PLIKSi) administered by trained psychology graduates. The PLIKSi consists of 12 core items derived from the Diagnostic Interview Schedule for Children, Version IV (DISC-IV) (Shaffer et al., 2000) and the Schedules for Clinical Assessment in Neuropsychiatry, Version 2.0 (SCAN 2.0) (World Health Organization Division of Mental Health, 1994). These items assess the presence, frequency and context of experiences associated with psychosis from three main domains of positive psychotic symptoms: hallucinations (visual and auditory), delusions (spied on, persecution, thoughts read, reference, control, grandiosity and other), and experiences of thought interference (broadcasting, insertion and withdrawal). Coding of all items followed glossary definitions, assessment, and rating rules for the SCAN 2.0. The PLIKSi administered at age 24 showed good inter-rater and test-retest reliability (intraclass correlation=0.81 [95% CI = 0.68-0.89] and intraclass correlation=0.90 [95% CI = 0.83-0.95]) (Sullivan et al., 2020).

Individuals who self-reported PEs were cross-questioned to verify the presence of each of these experiences, which were then rated by interviewers as: PEs not present, suspected PEs, or definite PEs. Cases of psychotic disorder were defined as having an interviewer-rated definite PE not attributable to the effects of sleep or fever, with recurring symptoms of at least once a month over the prior 6 months, which the individual reported as either very distressing, or had a negative impact on their social or occupational life, or led them to seek professional help (Zammit et al., 2013).

Negative symptoms such as apathy, anergia and asociality were assessed using 10 questions based on the Community Assessment of Psychic Experiences (CAPE) self-report questionnaire, where each item was rated on a 4-point scale (0=never; 1=sometimes; 2=often; 3=always) and summed. Missing items were handled with median replacement, except where all items were missing, the sum score was coded as missing. We then derived a binary variable based on the sum score, and we assigned those with a score of 14 or above into the ‘high negative symptoms’ group (Jones et al., 2016); this corresponded to 13.9% of the sample.

For the set of analyses looking at positive symptoms, the exposures were: i) ‘any PEs’: presence of any PEs, suspected or definite, not attributable to the effects of sleep or fever, ii) ‘definite PEs’: presence of any definite PEs not attributable to the effects of sleep or fever and iii) psychotic disorder as described above. The control group was defined as those who did not meet criteria for suspected/definite PEs or psychotic disorder. For the analysis looking at negative symptoms, the exposure was presence of high negative symptoms.

### 2.4. Blood inflammatory proteins at age 24 years

Blood samples were collected in lithium heparin tubes and placed on ice until processing. Samples were spun at 1300g for 10 minutes at 4-5°C, then plasma was aliquoted from the tube and stored at -80°C. Target time from sample collection until plasma frozen was 90 minutes. Heparin-stored plasma samples collected from at approximately age 24 (fasting) were analyzed using the Olink Target 96 Inflammation panel (Olink Analysis Service, Uppsala, Sweden). The Olink Target 96 Inflammation panel measures a selection of 92 proteins involved in inflammatory and immune response processes. Data were returned in Normalised Protein eXpression (NPX) values on a log_2_ scale, which are calculated from protein concentrations (cycle threshold [Ct] values) with normalization to minimize intra- and inter-assay variation (Olink Proteomics, 2021). Further details on the proteins assayed and quality control procedures applied have been previously reported (see data note by Goulding et al., 2024).

Samples that did not pass Olink’s quality control were excluded from the analysis. Proteins with ≥50% values below the limit of detection (LOD) were excluded (based on values reported in the relevant ALSPAC data note (Goulding et al., 2024); namely ARTN, Beta-NGF, FGF-23, FGF-5, GDNF, IL-1 alpha, IL-10RA, IL-13, IL-15RA, IL-17A, IL-2, IL-20, IL-20RA, IL-22 RA1, IL-24, IL-2RB, IL-33, IL-4, IL-5, LIF, NRT, NT-3, SIRT2, SLAMF1, TSLP), leaving 67 proteins to be used as outcomes in the analyses (**Table S1**). NPX values beyond 5 standard deviations (SD) from the mean were recoded as missing. No further normalization procedures were applied to the proteomic data prior to analysis, as it will reduce information available for the empirical Bayes estimation used in the analysis.

### 2.5. Covariables

BMI was computed using the formula: weight (kg) / height^2^ (m^2^), and we recoded values of <15 or >=60 as missing. Smoking was assessed via a questionnaire, and we derived a composite smoking variable by coding those who self-reported having smoked in the past 30 days and ≥1 cigarettes per day or ≥7 per week as smokers.

### 2.6. Statistical analysis

Sociodemographic characteristics of cases and controls were summarized as n (%), mean (standard deviation) or median [interquartile range] depending on the variable type (continuous or categorical). Group comparisons were conducted using Fisher’s exact tests for categorical variables, *t*-tests for normally distributed continuous variables and Kruskal-Wallis tests for non-normally distributed continuous variables.

Prior to analysis, correlations between inflammatory proteins were examined using Spearman’s rho (**Figure S1**) and a principal component analysis was performed to examine the overall distribution of the data and identify potential sample clustering in the inflammation proteomics panel. There was evidence for clustering effects by sex and BMI, but not by age, smoking, batch, or plate (**Figure S2**). Associations between each case definition and inflammatory proteins were evaluated using multiple linear models fitted in the *limma* R package (Ritchie et al., 2015), which uses an empirical Bayes method to moderate the standard errors of the estimated log-fold changes by accounting for information from other markers. For each of the case definitions, we ran a set of unadjusted models and a set of adjusted models with age, sex at birth, BMI and smoking as covariables. Correction for multiple comparisons was performed using the Benjamini-Hochberg procedure with a false discovery rate (FDR) adjusted *p*-value threshold of <0.05. We did not apply a fold change threshold as the effects are expected to be small in multifactorial phenotypes in a population-based sample.

We then examined protein-protein interactions (PPI) between the identified differentially abundant proteins at the nominal *p* < 0.05 level (separately for positive and negative symptoms) and functional enrichment in STRING database (version 12.0; https://string-db.org) (Szklarczyk et al., 2019). We used 66 out of the 67 proteins included in the analysis as the statistical background as STRING was not able to map VEGF-A to an entry in the database.

Data extraction and initial data cleaning was performed in StataMP 18 (StataCorp, 2023). Further data preparation and statistical analyses were conducted in RStudio 2022.07.1 using R v4.2.1 (R Core Team, 2022), using packages *tidyverse* (v2.0.0), *haven* (v2.5.2), *reshape2* (v1.4.4), *tableone* (v0.13.2), and *limma* (v3.54.2). Plots were generated using *ggplot2* (v3.4.2), *ggfortify* (v0.4.16), *ggrepel* (v0.9.3), and *ggpubr* (v0.6.0).

## 3. Results

### 3.1. Sample characteristics

Characteristics of the controls (n=2574) and any PEs (n=280), definite PEs (n=169), and psychotic disorder groups (n=67) at age 24 years from the ALSPAC birth cohort are presented in **Table 1**. Group comparisons revealed individuals in the psychotic disorder group were more likely to be female, those in the any PEs group were more likely to have higher BMI and all case groups were more likely to be smokers and report higher negative symptoms than controls. Comparisons between individuals with and without high negative symptoms showed that individuals with high negative symptoms (n=390) were marginally older, more likely to be smokers and less likely to have risky alcohol consumption than those with low or no negative symptoms (**Table 2**).

**Table 1.**
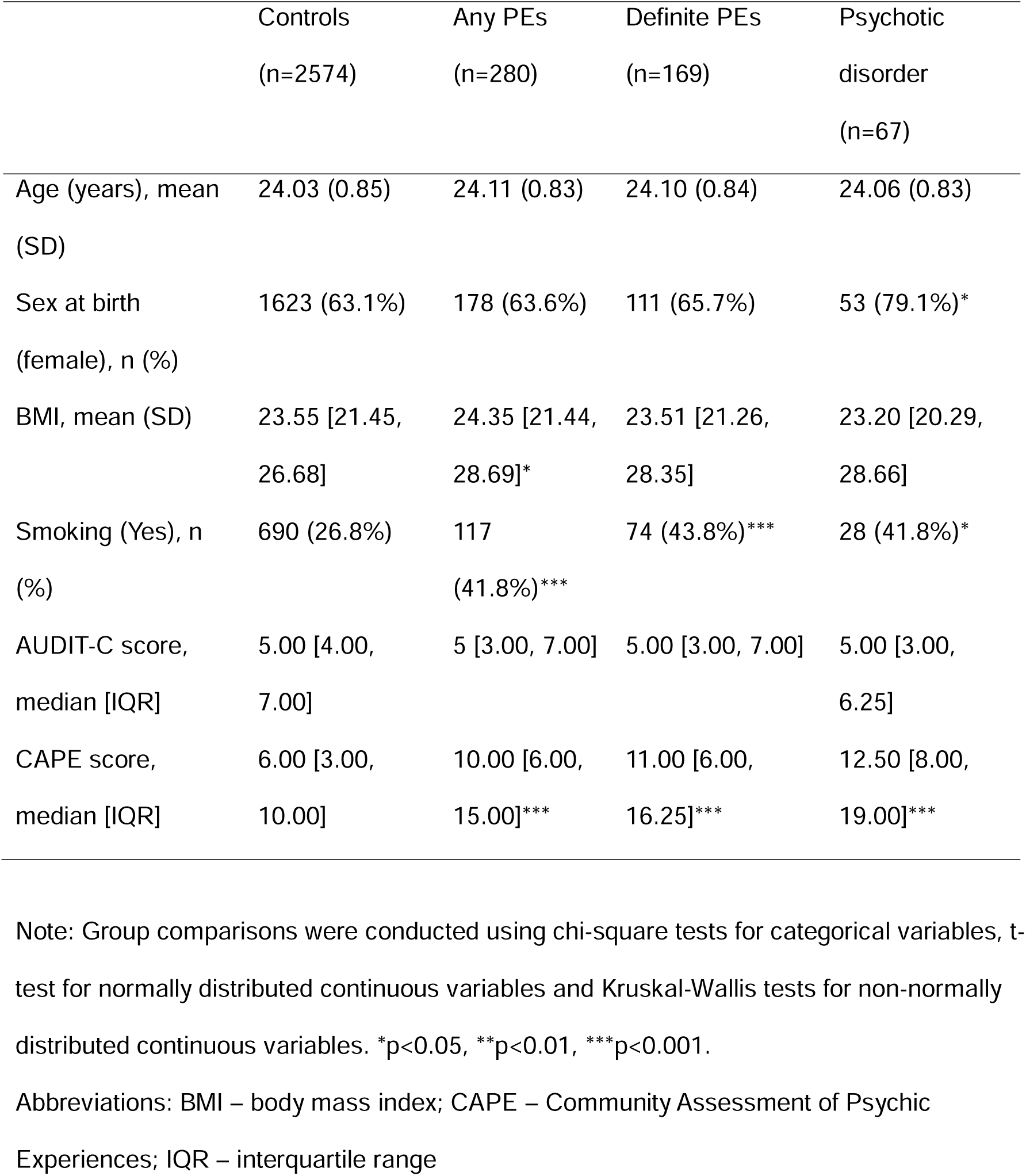
Sociodemographic and lifestyle characteristics of the included sample by psychosis outcome.

**Table 2.**
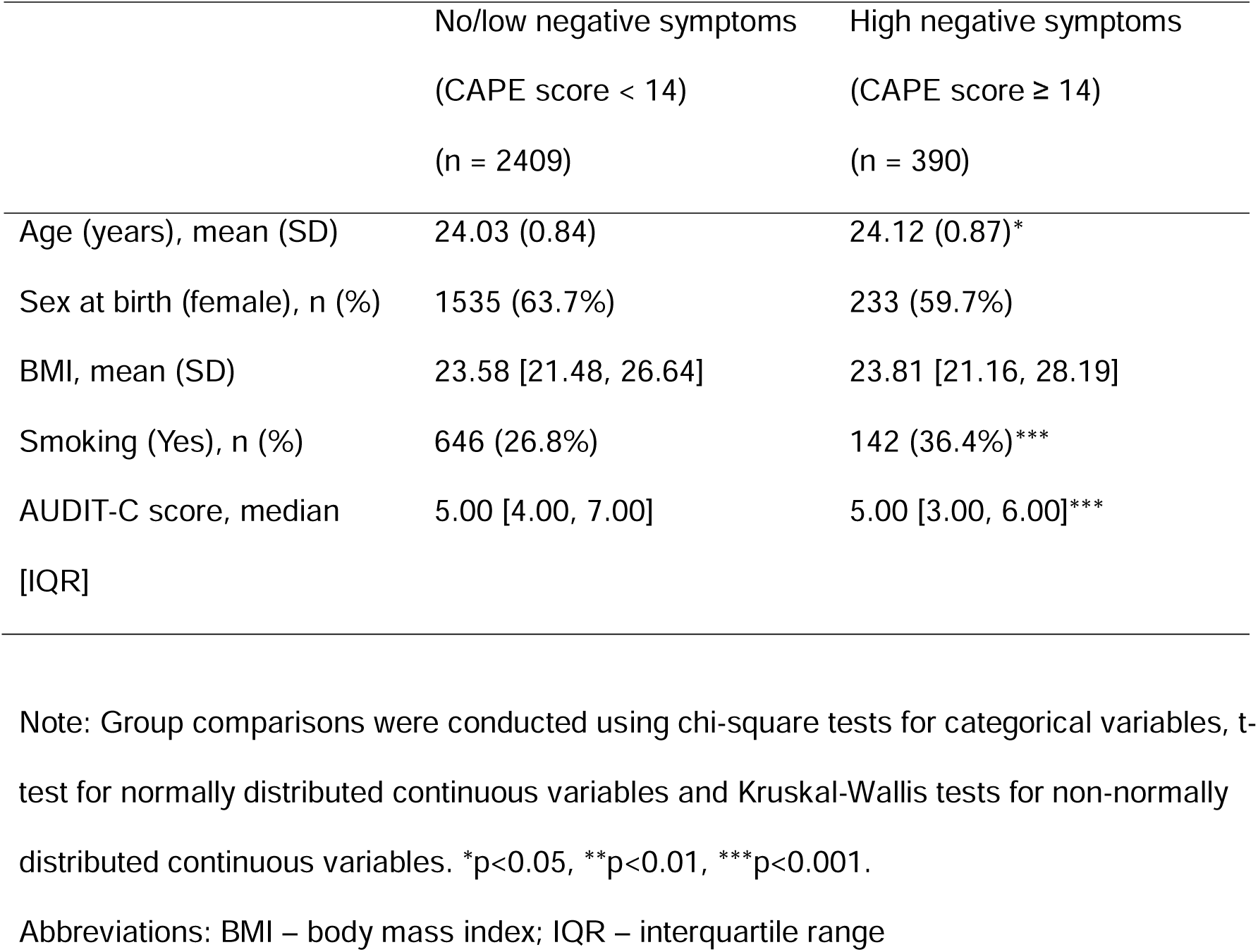
Sociodemographic and lifestyle characteristics by level of negative symptoms.

|  | No/low negative symptoms<br>(CAPE score < 14)<br>(n = 2409) | High negative symptoms<br>(CAPE score ≥ 14)<br>(n = 390) |
| --- | --- | --- |
| Age (years), mean (SD) | 24.03 (0.84) | 24.12 (0.87)* |
| Sex at birth (female), n (%) | 1535 (63.7%) | 233 (59.7%) |
| BMI, mean (SD) | 23.58 [21.48, 26.64] | 23.81 [21.16, 28.19] |
| Smoking (Yes), n (%) | 646 (26.8%) | 142 (36.4%)* |
| AUDIT-C score, median<br>[IQR] | 5.00 [4.00, 7.00] | 5.00 [3.00, 6.00]* |

Note: Group comparisons were conducted using chi-square tests for categorical variables, t- test for normally distributed continuous variables and Kruskal-Wallis tests for non-normally distributed continuous variables. \*p<0.05, \*\*p<0.01, \*\*\*p<0.001.
Abbreviations: BMI – body mass index; IQR – interquartile range

### 3.2. Differentially abundant inflammatory proteins in psychosis and related outcomes

The condition ‘any PEs’ was associated with upregulation of 22 inflammatory proteins after covariable adjustment; p-values of 6 of these associations survived FDR correction, which are CUB domain-containing protein 1 (CDCP1), TNF-related apoptosis-inducing ligand (TRAIL), IL-6, hepatocyte growth factor (HGF), monocyte chemoattractant protein-1 (MCP-1) and vascular endothelial growth factor A (VEGF-A) (**Figure 2B**). Definite PEs were associated with upregulation of 17 proteins after covariable adjustment, but only CDCP1 and IL-6 survived FDR correction (**Figure 2B**). Psychotic disorder was associated with upregulation of 11 proteins after covariable adjustment and evidence for three of these proteins, namely CDCP1, IL-6 and matrix metalloproteinase-10 (MMP-10), remained after FDR correction (**Figure 2B**). Full results for each of these models can be found in the **Supplementary Material**.

**Figure 2.**
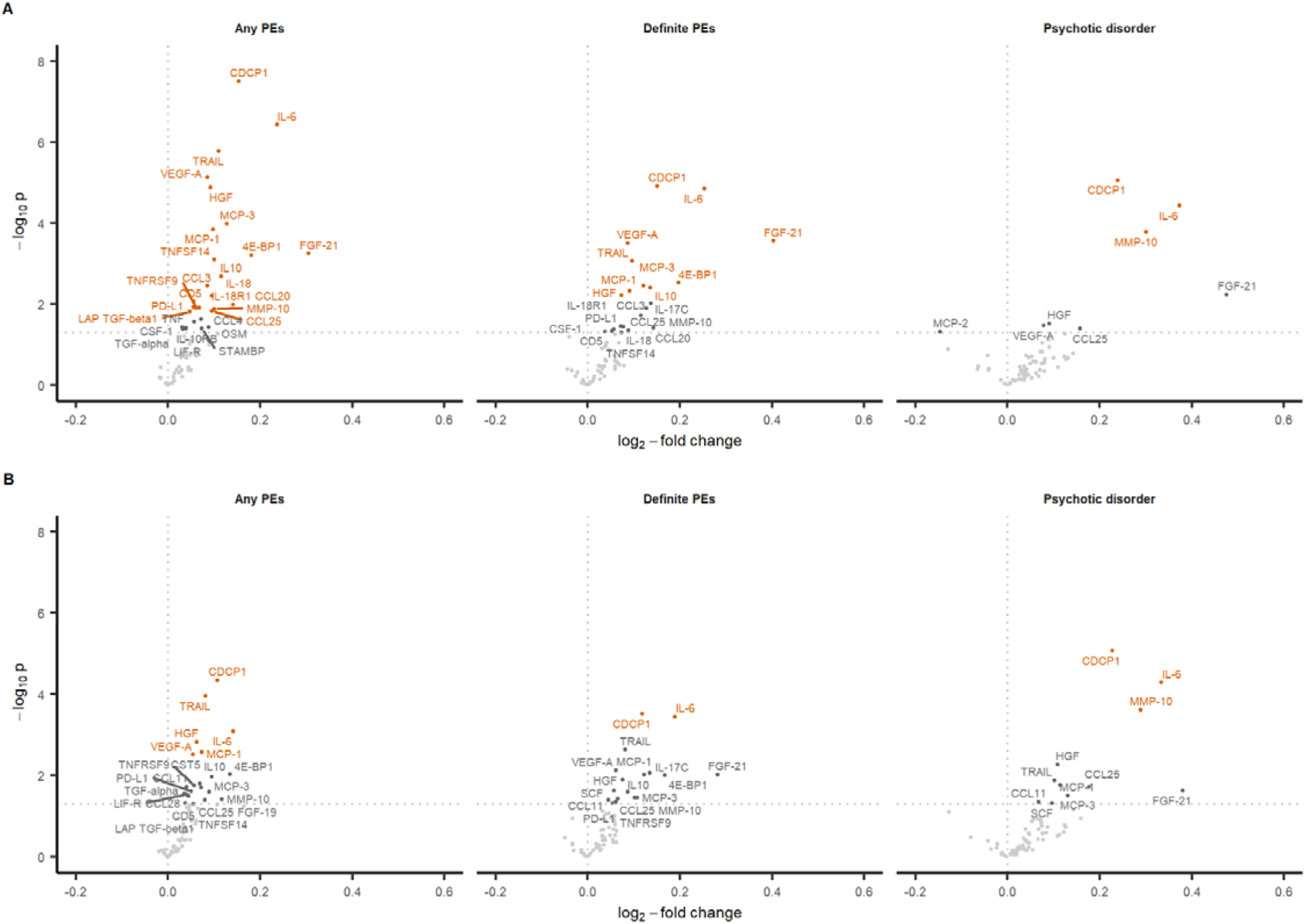
Differentially abundant blood inflammatory proteins associated with any psychotic experiences not attributable to sleep or fever, definite psychotic experiences not attributable to sleep or fever, and psychotic disorder. Panels A and B show results from unadjusted and adjusted models respectively. The labelled points are proteins that reach a nominal *p* < 0.05, and those in orange indicates upregulated proteins that survived FDR correction.

High negative symptoms were associated with upregulation of 23 proteins after covariable adjustment, 11 of which survived FDR correction. These proteins were HGF, MMP-10, IL-6, CDCP1, C-C motif chemokine ligand (CCL) 11, TRAIL, C-X3-C motif chemokine ligand 1 (CX3CL1), Fms-related receptor tyrosine kinase 3 ligand (FLT3L), fibroblast growth factor 21 (FGF-21), IL-10 receptor subunit beta (IL-10RB), CCL25 (**Table 3**). Full results can be found in the **Supplementary Material**.

**Table 3.**
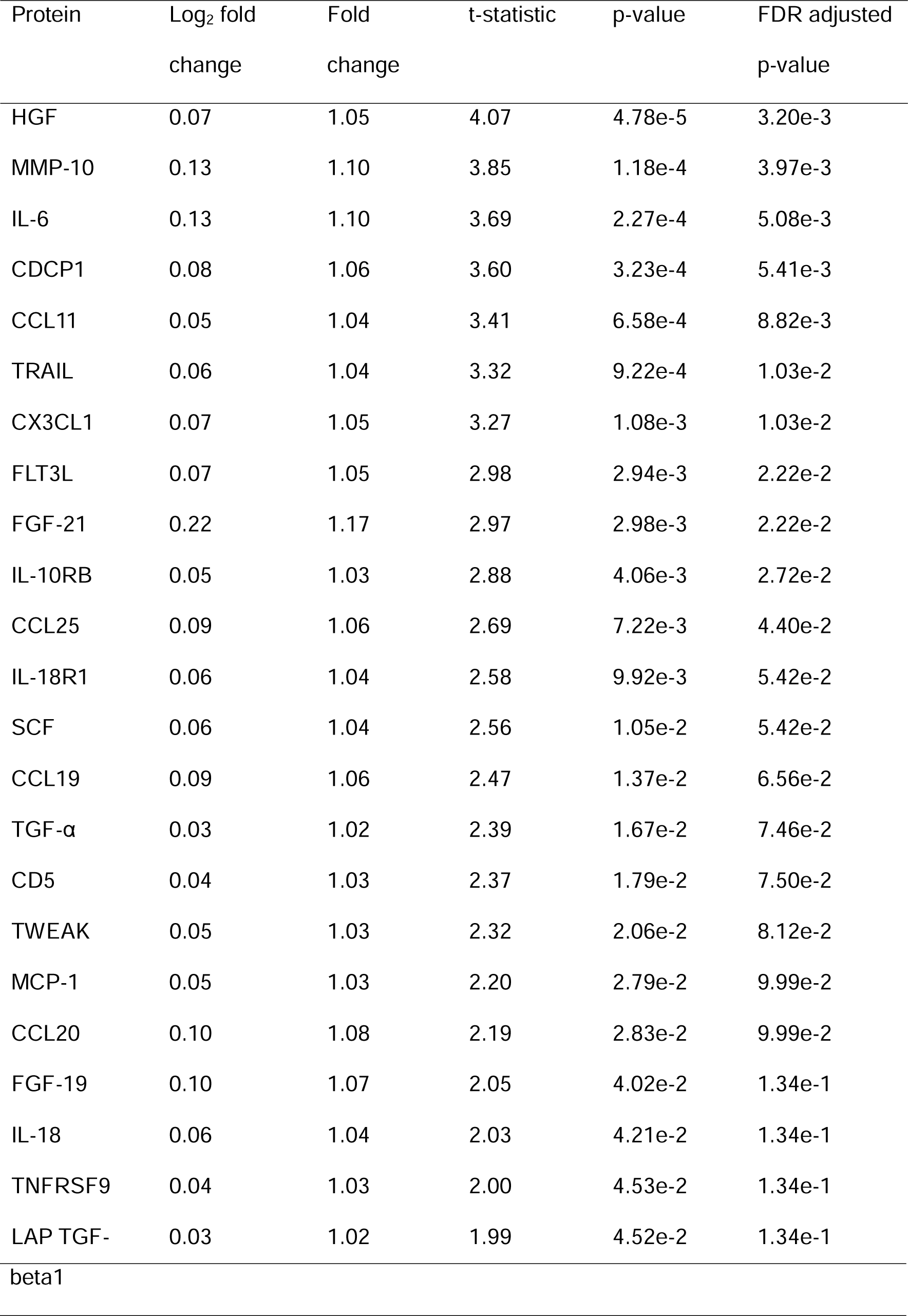
Inflammatory proteins associated with high negative symptoms, before and after FDR correction.

A total of 15 proteins showed upregulation in both positive and negative symptoms (using the nominal *p*-value cutoff of 0.05), which are CCL11, CCL25, T-cell surface glycoprotein CD5 (CD5), FGF-19, FGF-21, HGF, IL-6, latency-associated peptide transforming growth factor beta-1 (LAP TGF-β1), MCP-1, MMP-10, stem cell factor (SCF), transforming growth factor alpha (TGF-α), tumor necrosis factor receptor superfamily member 9 (TNFRSF9), and TRAIL.

### 3.4. Protein-protein interactions (PPI) and functional enrichment

Most identified differentially abundant proteins were functionally associated with each other within the protein-protein interaction network, suggesting they jointly contribute to shared functions (**Figures S3-S4**). However, the network showed no evidence of PPI or functional enrichment of Gene Ontology terms against the statistical background of the included list of proteins.

## 4. Discussion

Our study aimed to identify the plasma immune-inflammatory proteomic profile of psychotic disorder and related outcomes (positive and negative symptoms) in young adults using data from a well-phenotyped birth cohort. We observed upregulation of six, two and three proteins in any PEs, definite PEs and psychotic disorder respectively after covariable adjustment and FDR correction; CDCP1 and IL-6 were common to all three positive symptom case definitions and psychotic disorder was additionally associated with upregulation of MMP-10. We found upregulation of 11 protein in those with high negative symptoms, including CDCP1, IL-6 and MMP-10. These alterations were of relatively small magnitude, with many showing less than 10% upregulation, but more substantial fold changes were seen in the psychotic disorder group, where there were between 17% and 26% increases in CDCP1, IL-6 and MMP-10 levels. We observed alterations in different types of proteins, including growth factors, cytokines, chemokines, and matrix metalloproteinases that play roles in immune regulation, T cell activation and migration, macrophage activation and potentially blood-brain barrier (BBB) disruption, suggesting widespread immunological alterations are involved in psychosis.

We report consistent evidence for associations of IL-6 across all psychosis-related outcomes examined including positive, negative symptoms and psychotic disorder. IL-6 is a pleiotropic cytokine that plays a key role in the regulation of immune responses and inflammation. As the most frequently studied cytokine, there is considerable evidence that suggests circulating concentrations of IL-6 is elevated across the psychosis spectrum, including in those at risk of psychosis (Misiak et al., 2021), in antipsychotic-naïve FEP (Dunleavy et al., 2022) and in both acute and chronic schizophrenia spectrum disorder (Halstead et al., 2023). IL-6 has also been reported to be associated with negative symptoms (Dunleavy et al., 2022). Our findings support an ongoing clinical trial investigating whether inhibition of IL-6 signaling using an anti-IL6R monoclonal antibody, tocilizumab, improves negative symptoms in people with psychosis (Foley et al., 2023). Due to the pleiotropic effects of IL-6, the specific mechanisms underlying the associations between increased IL-6 and psychosis remain poorly understood and further research is required to elucidate the specific biological pathways and cascades involved.

CDCP1 is a transmembrane glycoprotein with three extracellular CUB domains. It has typically been studied in the context of cancer due to its overexpression in solid tumors and is considered a key regulator of cancer metastasis through its roles in cell migration and invasion, and extracellular matrix degradation via MMP-9 secretion (Miyazawa et al., 2010). More recently, CDCP1 has been found to have immune regulatory roles; it is a ligand for CD6 and regulates T cell activation and migration (Enyindah-Asonye et al., 2017), and reduced IL-6 production was observed in CDCP1 knockout mice in an animal model of Kawasaki disease (Lun et al., 2021). CDCP1 has been shown to participate in a CD6 tripartite receptor complex that induces cytoskeleton remodeling and cell barrier tight junction reduction in retinal pigment epithelium cells (Borjini et al., 2023), suggesting this process may underlie disruptions to tissue barriers caused by T cell-mediated inflammation. It is possible that this process also contributes to disrupted BBB and T cell infiltration into the central nervous system, leading to neuroinflammation and neuropsychiatric disorders such as psychosis and schizophrenia spectrum disorders.

MMPs are a family of zinc-dependent endopeptidases that contribute to tissue remodeling by degradation of extracellular matrix components in normal physiological processes and pathological conditions. In recent years, MMP-9 emerged as a candidate involved the pathophysiology of schizophrenia, as it is known to regulate a range of neurodevelopmental processes including synaptic and dendritic spine plasticity, neuroinflammation and BBB permeability (Bitanihirwe and Woo, 2020; Chopra et al., 2019). Although studies examining the candidate functional single nucleotide polymorphism (SNP) -1562 C/T polymorphism (rs3918242) in the *MMP9* gene reported conflicting results (Xia et al., 2019), there is evidence for peripheral MMP-9 upregulation in schizophrenia spectrum disorders (Schoretsanitis et al., 2021).

MMP-10, also known as stromelysin-2, is expressed by macrophages in response to injury, infection or transformation; it is a critical mediator of macrophage activation (McMahan et al., 2016) and has been shown to promote chronic inflammation (Sánchez et al., 2022). MMP-10 has rarely been studied in the context of neuropsychiatric disorders, and the limited literature has reported mixed findings in relation to schizophrenia. One study reported that elevated CSF MMP-10 levels were associated with major depressive disorder, but not schizophrenia (Omori et al., 2020); however, another study reported associations between epigenetic proxies of plasma concentrations of MMP-10 and clinical features of schizophrenia (Kiltschewskij et al., 2024). It has also been shown *in vitro* that MMP-10 can activate MMP-9 via proteolysis of its precursor pro-MMP9 (Nakamura et al., 1998) and thus may be involved in a cascade that impacts on neurodevelopment and neuroinflammation in psychosis and schizophrenia spectrum disorders.

The novel markers of CDCP1 and MMP-10 suggest potential roles of BBB dysfunction and extracellular matrix abnormalities in the pathophysiology of psychosis. Neuroinflammation has been shown to contribute to neurovascular endothelial dysfunction and increased BBB permeability in schizophrenia (Najjar et al., 2017). Extracellular matrix abnormalities, including reduced perineuronal nets and reduced extracellular matrix gene expression in several brain regions (Pantazopoulos et al., 2021) and alterations in brain extracellular matrix composition (Rodrigues-Amorim et al., 2022) have also been reported in schizophrenia. Further research is required to better understand the cascade of events including disruption of BBB integrity and extracellular matrix abnormalities in the pathogenesis of psychosis.

As a result of the cross-sectional design used here, we cannot ascertain whether any of the protein associations are causal for psychosis. However, we looked up evidence of potential causality based on existing Mendelian randomization (MR) studies which use genetic variants as instrumental variables to assess whether the association between an exposure and an outcome is likely to be causal or result of residual confounding or reverse causation (Sanderson et al., 2022). Existing MR studies support a potential causal role of IL-6 in schizophrenia (Hartwig et al., 2017; Perry et al., 2021), with also weak evidence of MCP-1 (Perry et al., 2021) that was associated with both positive and negative symptoms in the present study. In addition, a more recent MR study involving 735 immune-related biomarkers reported MR evidence for potential causal effects with schizophrenia for several proteins we found to be associated with psychosis/psychotic symptoms in our sample. These include IL-6, IL-10RB, IL-18, LIF-R, CCL25, FGF-19, and MMP-10 (Dardani et al., 2024). In particular, the study by Dardani and colleagues reported stronger evidence of causality for FGF-19 and MMP-10, though the interpretation may be complicated due to the use of potentially complex genetic instruments derived from trans quantitative trait loci (trans-QTLs) associations. In addition, FGF19 also showed evidence of genetic colocalization whereby the locus associated with blood FGF-19 protein levels were also associated with schizophrenia (Zuber et al., 2022). Taken together, findings from our analysis reported here and existing MR studies suggest that FGF-19 and MMP-10 could be causally relevant for schizophrenia. These proteins (along with others highlighted by our study and existing MR studies) require further investigation.

Strengths of this study include using detailed blood immune proteomic data, various psychosis-related outcomes across the psychosis spectrum as well as examining both positive and negative symptoms, as well as using a population-based young adult sample. There are several limitations to the existing study. Firstly, this study is cross-sectional in design, making it difficult to ascertain whether any observed associations are causal or result of reverse causality or residual confounding. Further research is required to replicate these associations, determine the temporal order of events and investigate whether these associations are causal. Secondly, as the study sample is drawn from a population-based cohort, there is likely a larger degree of heterogeneity in symptom severity, phase, duration, psychotropic medication use and neuropsychiatric comorbidity within the case groups, for which we have not been able to adjust despite their potential effects on blood protein levels. Lastly, the case groups defined by positive symptoms are rather small and the analyses may be underpowered to detect smaller effects, which may explain why only few signals were found relative to the analysis involving negative symptoms.

## 5. Conclusions

Using data from a well characterized birth cohort, we report that IL-6 and CDCP1 are associated with psychotic disorder and positive symptoms of psychosis, while negative symptoms are associated with changes in a larger number of immune proteins which partly overlap with proteins associated with positive symptoms. In addition to IL-6 that is already well reported in the literature, we identified two novel inflammatory markers, CDCP1 and MMP-10, which are implicated in immune regulation, T cell activation and migration, macrophage activation, BBB disruption, and extracellular matrix abnormalities. Our findings suggest there are widespread alterations in different types of inflammatory markers in young people with psychosis or psychotic symptoms. This is consistent with a role of inflammation and immune dysfunction in the pathogenesis of psychotic disorders.

## Data Availability

ALSPAC data access is through a system of managed open access. Information on how to request access to ALSPAC data can be found on the ALSPAC website (http://www.bristol.ac.uk/alspac/researchers/access/).

## CRediT authorship contribution statement

Ruby Tsang: Conceptualization, Formal analysis, Writing – original draft, Writing – review & editing

Nicholas Timpson: Conceptualization, Writing – review & editing, Supervision

Golam Khandaker: Conceptualization, Writing – review & editing, Supervision

## Acknowledgements

We are extremely grateful to all the families who took part in this study, the midwives for their help in recruiting them, and the whole ALSPAC team, which includes interviewers, computer and laboratory technicians, clerical workers, research scientists, volunteers, managers, receptionists and nurses.

