## Supplementary Material for "Inflammation Proteomic Profiling of Psychosis in Young Adults: findings from the ALSPAC birth cohort"

Figure S1. Heatmap of Spearman’s correlations with hierarchical clustering between included immune-inflammatory proteins for the included sample.

Figure S2. Principal component plots to assess potential clustering effects of covariables.

Figure S3. STRING-DB plot of protein-protein interactions for positive symptoms.

Figure S4. STRING-DB plot of protein-protein interactions for negative symptoms.

Table S1. List of 67 analyzed inflammation proteomic markers.

Table S2. Results for the unadjusted and adjusted models for any psychotic experiences.

Table S3. Results for the unadjusted and adjusted models for definite psychotic experiences.

Table S4. Results for the unadjusted and adjusted models for psychotic disorder.

Table S5. Results for the unadjusted and adjusted models for negative symptoms.

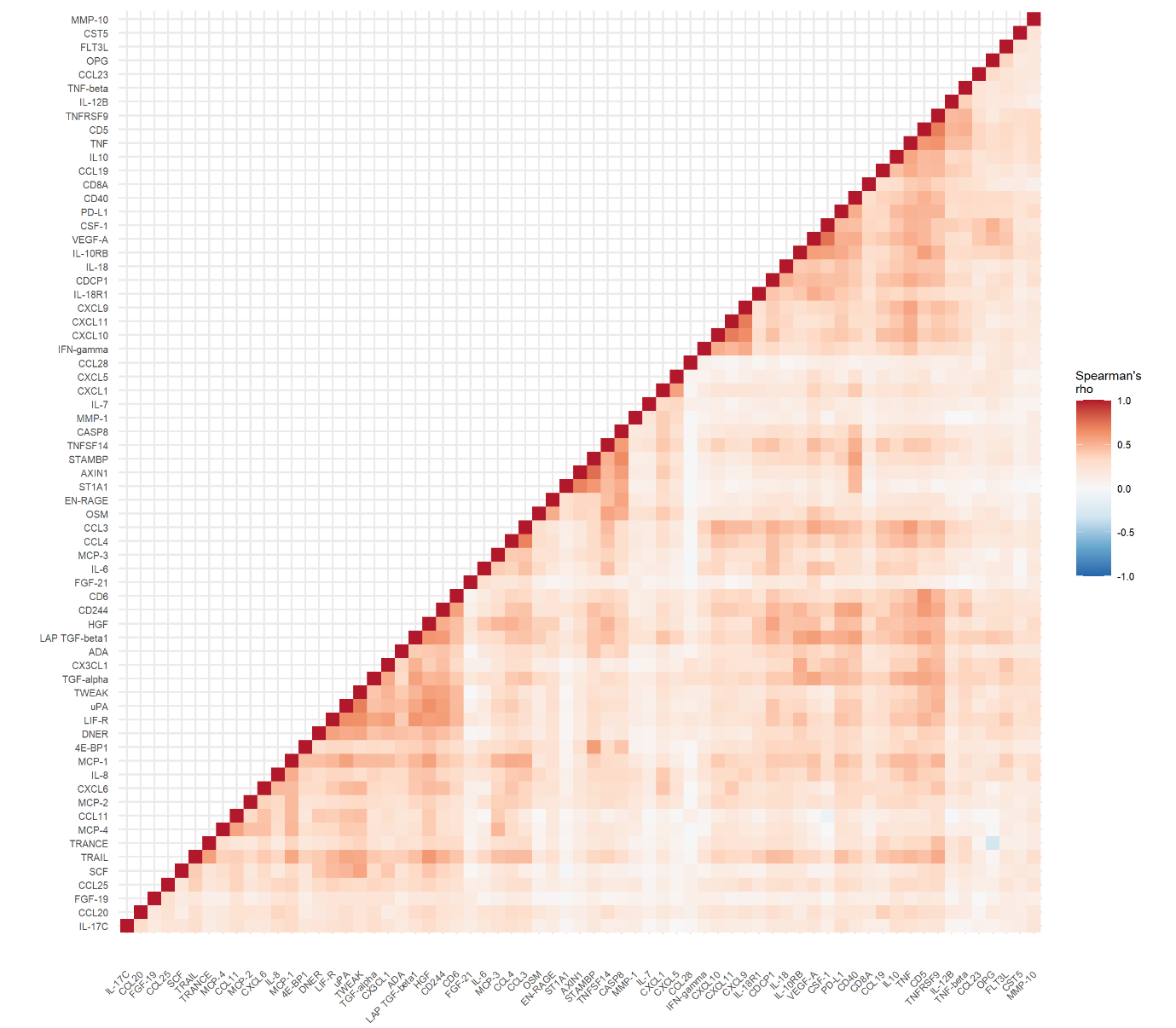

**Figure S1. Heatmap of Spearman’s correlations with hierarchical clustering between included immune-inflammatory proteins for the included sample.**

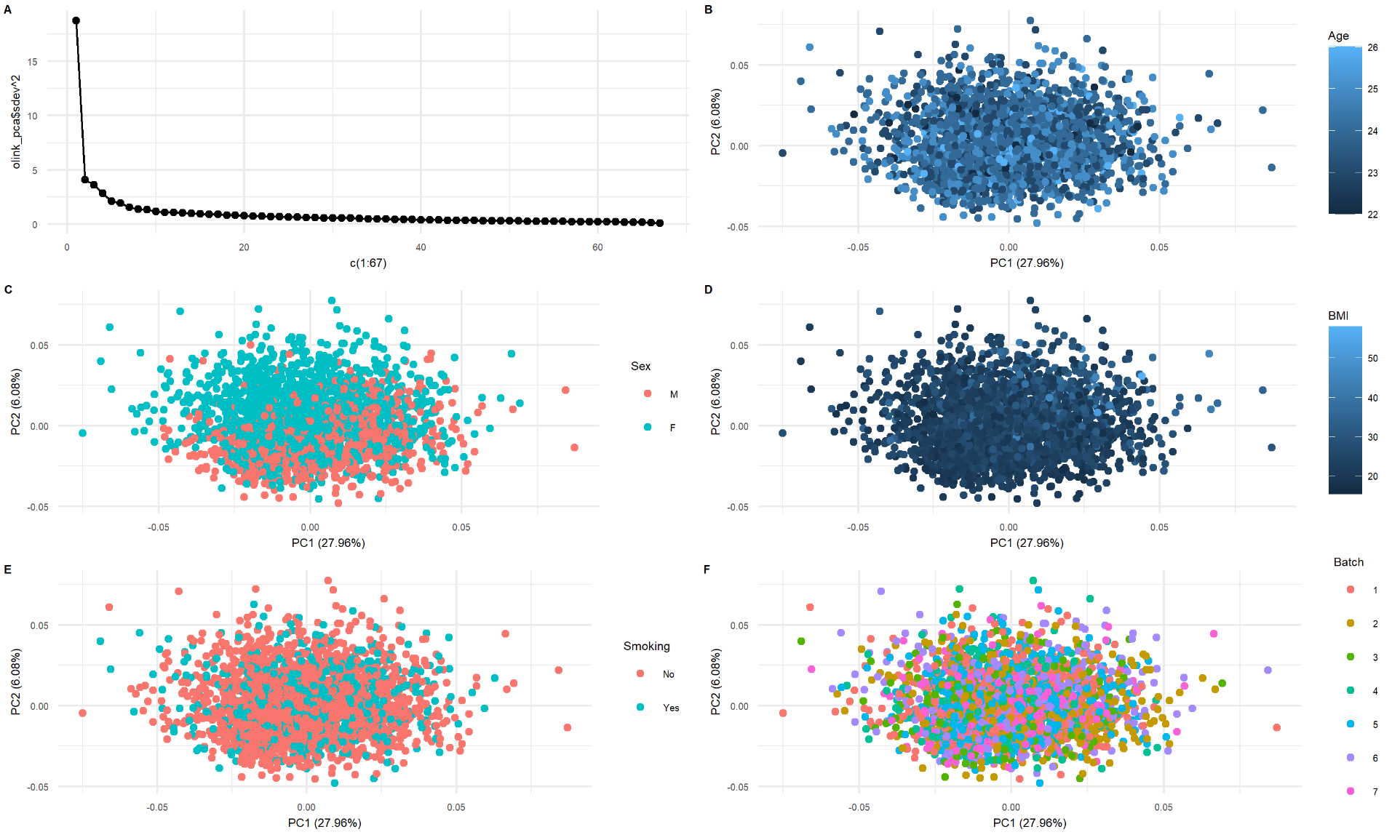

**Figure S2. Scree plot of the principal components analysis, and principal component plots to assess potential clustering effects of covariables.**

Panel A shows the scree plot of the proteomics data, and panels B to F are plots assessing potential clustering effects of age, sex, BMI, smoking and batch.

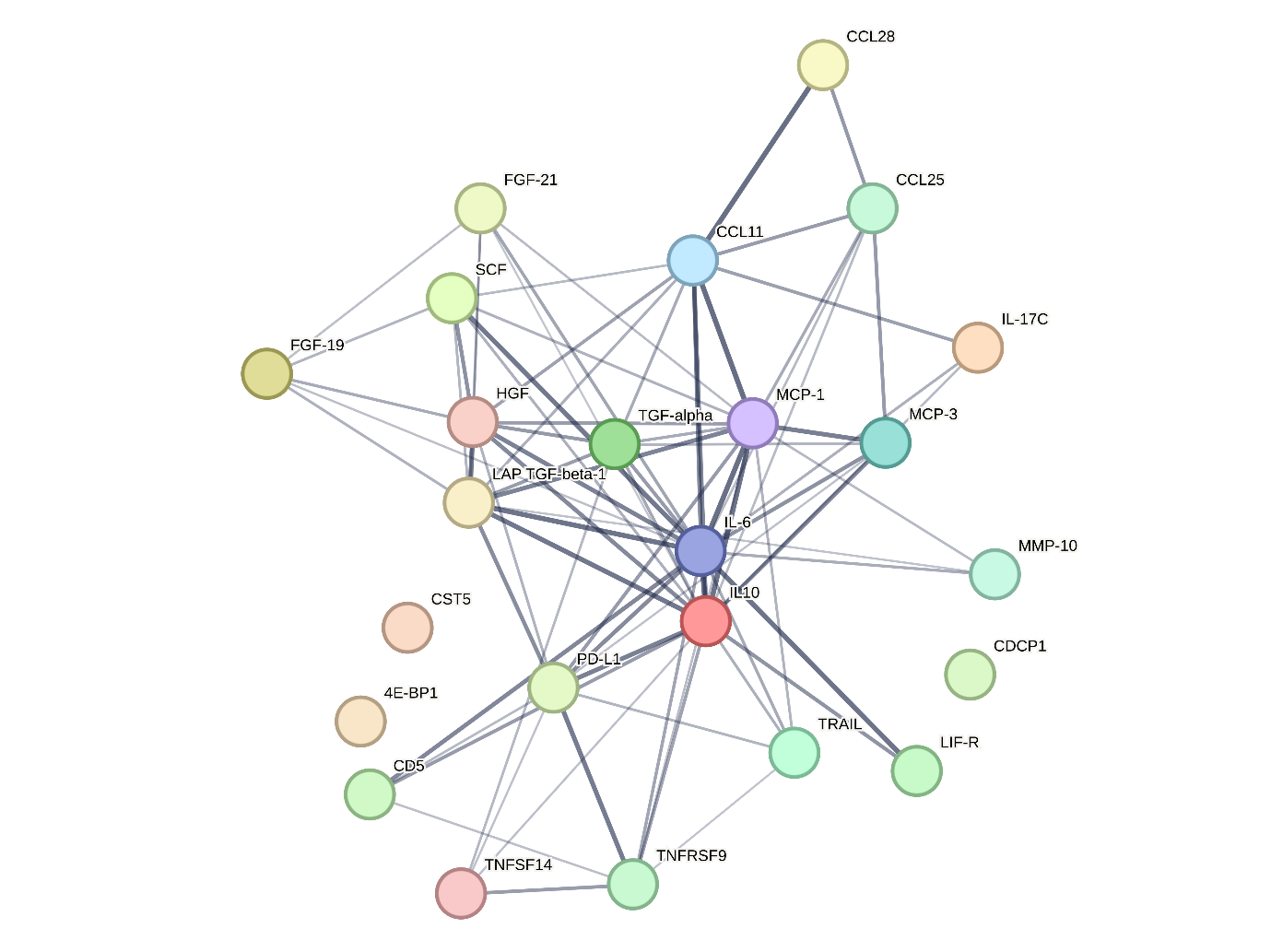

**Figure S3. STRING-DB plot of protein-protein interactions for positive symptoms.**

Colored nodes represent proteins that were entered into the query. Edges represent protein-protein associations (i.e. they jointly contribute to a shared function), where the line thickness indicates the strength of support for the association.

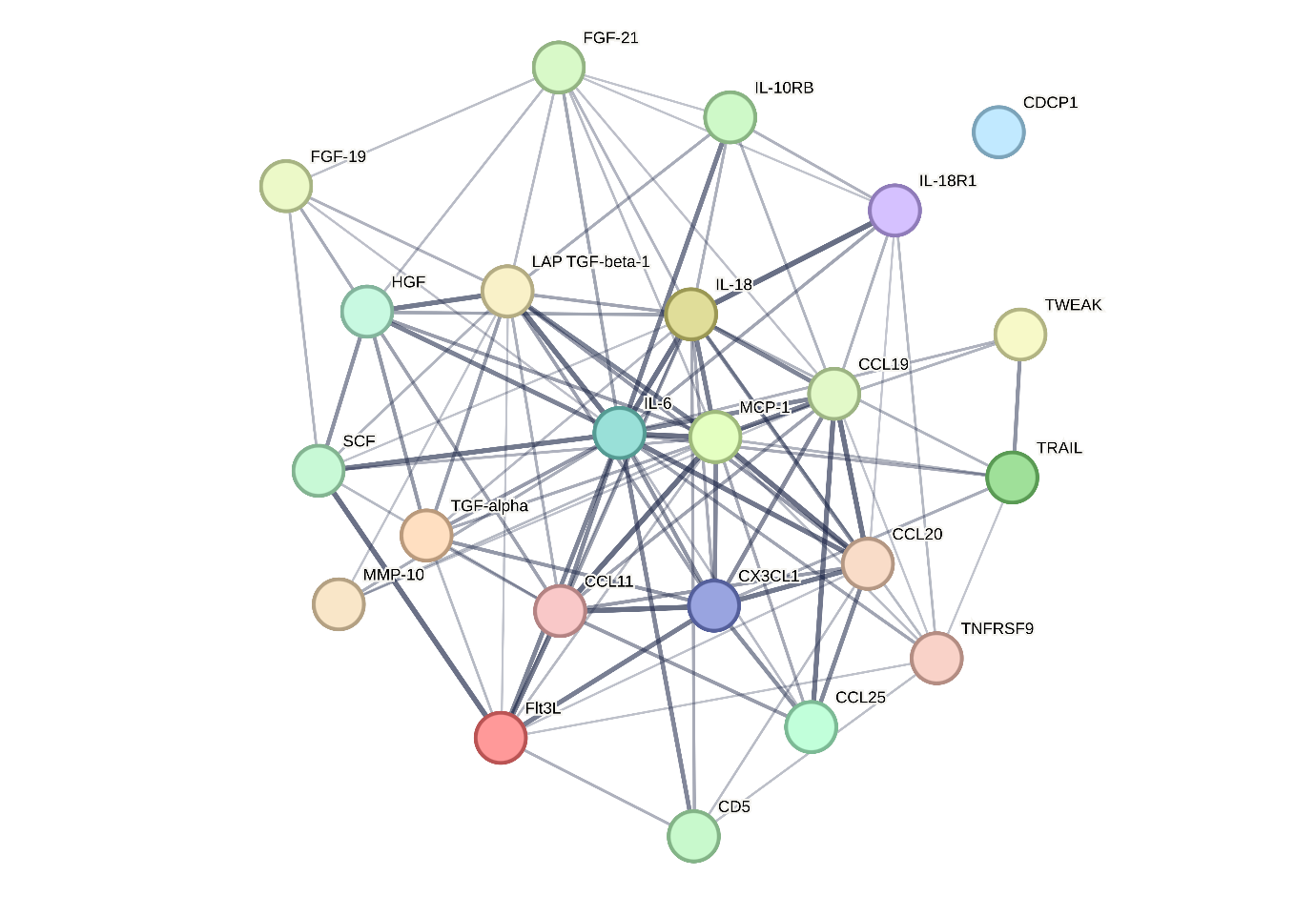

**Figure S4. STRING-DB plot of protein-protein interactions for negative symptoms.**

Colored nodes represent proteins that were entered into the query. Edges represent protein-protein associations (i.e. they jointly contribute to a shared function), where the line thickness indicates the strength of support for the association.

**Table S1. List of 67 analyzed inflammation proteomic markers.**

| Full name | Short name | UniprotID |
| --- | --- | --- |
| Eukaryotic translation initiation factor 4E-binding protein 1 | 4E-BP1 | Q13541 |
| Adenosine Deaminase | ADA | P00813 |
| Axin-1 | AXIN1 | O15169 |
| Caspase-8 | CASP-8 | Q14790 |
| Eotaxin | CCL11 | P51671 |
| C-C motif chemokine 19 | CCL19 | Q99731 |
| C-C motif chemokine 20 | CCL20 | P78556 |
| C-C motif chemokine 23 | CCL23 | P55773 |
| C-C motif chemokine 25 | CCL25 | O15444 |
| C-C motif chemokine 28 | CCL28 | Q9NRJ3 |
| C-C motif chemokine 3 | CCL3 | P10147 |
| C-C motif chemokine 4 | CCL4 | P13236 |
| Natural killer cell receptor 2B4 | CD244 | Q9BZW8 |
| CD40L receptor | CD40 | P25942 |
| T-cell surface glycoprotein CD5 | CD5 | P06127 |
| T cell surface glycoprotein CD6 isoform | CD6 | P30203 |
| T-cell surface glycoprotein CD8 alpha chanin | CD8A | P01732 |
| CUB domain-containing protein 1 | CDCP1 | Q9H5V8 |
| Macrophage colony-stimulating factor 1 | CSF-1 | P09603 |
| Cystatin D | CST5 | P28325 |
| Fractalkine | CX3CL1 | P78423 |
| C-X-C motif chemokine 1 | CXCL1 | P09341 |
| C-X-C motif chemokine 10 | CXCL10 | P02778 |
| C-X-C motif chemokine 11 | CXCL11 | O14625 |
| C-X-C motif chemokine 5 | CXCL5 | P42830 |
| C-X-C motif chemokine 6 | CXCL6 | P80162 |
| C-X-C motif chemokine 9 | CXCL9 | Q07325 |
| Delta and Notch-like epidermal growth factor-related receptor | DNER | Q8NFT8 |
| Protein S100-A12 | EN-RAGE | P80511 |
| Fibroblast growth factor 19 | FGF-19 | O95750 |
| Fibroblast growth factor 21 | FGF-21 | Q9NSA1 |
| Fms-related tyrosine kinase 3 ligand | Flt3L | P49771 |
| Hepatocyte growth factor | HGF | P14210 |
| Interferon gamma | IFN-gamma | P01579 |
| Interleukin-10 receptor subunit beta | IL-10RB | Q08334 |
| Interleukin-12 subunit beta | IL-12B | P29460 |
| Interleukin-17C | IL-17C | Q9P0M4 |
| Interleukin-18 | IL-18 | Q14116 |
| Interleukin-18 receptor 1 | IL-18R1 | Q13478 |
| Interleukin-6 | IL-6 | P05231 |
| Interleukin-7 | IL-7 | P13232 |
| Interleukin-8 | IL-8 | P10145 |
| Interleukin-10 | IL10 | P22301 |
| Latency-associated peptide transforming growth factor beta-1 | LAP TGF-beta-1 | P01137 |
| Leukemia inhibitory factor receptor | LIF-R | P42702 |
| Monocyte chemotactic protein 1 | MCP-1 | P13500 |
| Monocyte chemotactic protein 2 | MCP-2 | P80075 |
| Monocyte chemotactic protein 3 | MCP-3 | P80098 |
| Monocyte chemotactic protein 4 | MCP-4 | Q99616 |
| Matrix metalloproteinase-1 | MMP-1 | P03956 |
| Matrix metalloproteinase-10 | MMP-10 | P09238 |
| Osteoprotegerin | OPG | O00300 |
| Oncostatin-M | OSM | P13725 |
| Programmed cell death ligand 1 | PD-L1 | Q9NZQ7 |
| Stem cell factor | SCF | P21583 |
| Sulfotransferase 1A1 | ST1A1 | P50225 |
| STAM-binding protein | STAMBP | O95630 |
| Transforming growth factor alpha | TGF-alpha | P01135 |
| Tumor necrosis factor | TNF | P01375 |
| TNF-beta | TNFB | P01374 |
| Tumor necrosis factor receptor superfamily member 9 | TNFRSF9 | Q07011 |
| Tumor necrosis factor ligand superfamily member 14 | TNFSF14 | O43557 |
| TNF-related apoptosis-inducing ligand | TRAIL | P50591 |
| TNF-related activation-induced cytokine | TRANCE | O14788 |
| Tumor necrosis factor (Ligand) superfamily, member 12 | TWEAK | O43508 |
| Urokinase-type plasminogen activator | uPA | P00749 |
| Vascular endothelial growth factor A | VEGF-A | P15692 |
